# Effects of a suspension training warm□up on cardiopulmonary exercise performance in recreationally active college□aged adults: a randomized crossover study protocol

**DOI:** 10.1101/2025.11.16.25340334

**Authors:** Roberto Federico, Tiffany A. Caba Abreu, Jamie Pigman, Tamara Rial Faigenbaum

**Affiliations:** Department of Health & Physical Education, Monmouth University, West Long Branch, NJ, USA

## Abstract

**Objectives:** This study will aim to assess the acute cardiopulmonary differences of a treadmill walking warm□up (WW) versus a suspension warm□up (SW) immediately preceding cardiopulmonary exercise testing (CPET) in recreationally active college□aged adults. The primary outcomes will be to assess differences in peak oxygen consumption, peak heart rate, and peak minute ventilation during CPET following a WW versus a SW. Secondary outcomes will include time to exhaustion, rate of perceived exertion, blood pressure, tidal volume, fraction of expired oxygen consumption, and respiration rate.

**Methods:** This study will be a randomized counterbalanced crossover design. Participants will complete two separate CPETs over two non□consecutive test days (≥48□h, ≤7□d). During each visit, participants will complete either a WW (6□min at self□selected pace on a treadmill, 0% grade) or SW (6□min dynamic sequence of suspension training exercises), followed by an incremental treadmill CPET protocol up to maximal exertion. Gas exchange, heart rate, blood pressure, and rate of perceived exertion will be measured at rest, after warm-up, during CPET protocol, immediately after, and after 5 minutes of resting. Analyses will use linear mixed-effects models and two one-sided tests for equivalence.

**Conclusions:** This protocol will determine whether a brief, dynamic suspension training warm□up is a practical and transferable approach or rather different from a traditional treadmill warm-up before CPET in active young adults.

**Ethics/registration:** IRB SP2553 (June 25, 2025), clinical trials Identifier: NCT07215052.

## BACKGROUND

Cardiopulmonary exercise testing (CPET) is commonly used in clinical and research settings to assess health status and aerobic fitness. Physiological responses to submaximal and maximal aerobic exercise can provide important information about cardiopulmonary function, abnormal exercise responses, and disease severity. In addition, CPET can assist in establishing a baseline biometric, designing an exercise program, tracking progress, and encouraging ongoing participation in physical activities. CPET is considered the gold standard because it combines standard graded exercise testing with simultaneous ventilatory respired gas analysis.

Dynamic warm□ups that integrate multi□joint movements raise core temperature, increase heart rate and ventilation, and can effectively prime oxygen□uptake kinetics; they are practical alternatives to simple walking or cycling before graded exercise testing. Suspension training uses bodyweight-based training through an adjustable strap system to perform a variety of exercises in an unstable environment. Suspension training is commonly performed with a total resistance exercise (TRX) training strap (www.trxtraining.com). Suspension training is widely used in fitness and rehabilitation settings due to its versatility, portability, and adaptability for individuals of any age and physical condition levels. Previous studies have used suspension training as a conditioning method for enhancing core stability, balance, and neuromuscular control (Smith et al., 2016; Aguilera-Castells et al., 2020).

While suspension training is commonly used in fitness and rehabilitation programs, further research is required to understand its adaptability and transferability to be a warm-up strategy alternative to traditional aerobic-based warm-ups before CPET. We are unaware of previous studies investigating the acute cardiometabolic and pulmonary responses to a suspension training warm-up protocol (SW). Smith et al. (2016) evaluated the acute cardiometabolic response to a suspension training conditioning session. However, this study assessed adults with low levels of exercise experience during a complete training session for conditioning purposes. Young, active, or athletic individuals may require different intensities and respond differently to specific warm-up exercise programs before CPET. Moreover, no known studies have evaluated suspension training as a viable warm-up option for CPET. To better understand adequate loads and program design for highly trained individuals, we propose to evaluate a brief dynamic SW protocol in young recreationally active adults. We believe that a dynamic SW will elicit an equivalent physiological response as a traditional continuous walking warm-up (WW) before CPET.

## OBJECTIVES

The primary objective of this study is to compare peak VO_2_ (L/min), minute ventilation (L/min) (V_E_), and HR peak during CPET following a SW or WW protocol. The secondary objectives of this study are to compare time to exhaustion, rate of perceived exertion (RPE), blood pressure (BP), tidal volume, fraction of expired oxygen consumption (FeO_2_), and respiration rate during CPET between the two warm-up protocols.

## METHODS

### Study design

This study is a randomized, counterbalanced two□condition crossover employing a Latin□square order. Two visits are scheduled ≥48□h and ≤7□d apart, at the same time of day under controlled environmental conditions. Estimation of sample size was done with an a priori power analysis run by the software G-Power (version 3.1.9, Heinrich-Heine-University, Düsseldorf, Germany), with a desired power level of 0.80, an alpha level of 0.05, and a large effect size (Cohen’s *d* = 0.8) using a two-tailed paired samples t-test (Faul et al., 2009). A total of 15 participants was calculated for a total power of 0.82.

### Participants

Inclusion criteria for participants include: a) between 19 to 25 years of age; b) accumulate a minimum of 5 hours per week of moderate to vigorous physical activity measured by the IPAQ questionnaire (Craig et al., 2003); c) be able to participate in moderate to vigorous physical activity as determined by the Physical Activity Readiness Questionnaire (PARQ, Warburton et al., 2023). Exclusion criteria are as follows: a) accumulate less than 5 hours per week of moderate or vigorous physical activity; b) have a health condition that is a contraindication for performing maximal CPET including cardiovascular disease, pulmonary issues, metabolic conditions, neuromuscular disorders, orthopedic limitations, or acute illness that impedes performing CPET and suspension exercise; d) not completing the study procedures or not providing informed consent and; e) not be able to participate in moderate to vigorous physical activity as determined by the PARQ-2023 questionnaire.

Before the start of the study, all participants will be informed about the study’s aims and procedures and will sign the informed consent form. This study was approved by the Institutional Review Board at Monmouth University. All participants will sign a consent form verifying that they meet the inclusion criteria and understand the benefits and risks of the study. All collected data will be deidentified. Data will be sourced and gathered respectfully and inclusively, adhering strictly to the inclusion and exclusion criteria listed above and following ethical principles outlined in the Declaration of Helsinki.

### Experimental Design

Participants will report to the Human Performance Laboratory of Monmouth University from the Department of Health and Physical Education situated at the Monmouth University Graduate Center, room 222 (West Long Branch, NJ). They will come on two nonconsecutive days (within 1 week maximum) at the same time of day and under the same controlled temperature and humidity conditions. Participants will be instructed not to participate in vigorous exercise within the 48 hours before the testing and to refrain from caffeine and alcohol consumption before the day of testing, as well as to avoid eating the 3 hours before the testing and to use the same shoes for both trial sessions. A Latin-square arrangement design will be used for the randomization of the session. Participants will be given standardized instructions before the start of the test and the same verbal encouragement during both testing procedures. All testing protocols will be supervised by the researchers of this study (TRR, JP) with experience in conducting and assessing CPET. The research team comprises two women and two men, including graduate and undergraduate students and researchers from various specialties and ethnic backgrounds.

The treadmill protocol test to be used is based on the protocol proposed by Green et al. (2023), which is a valid and reproducible exercise protocol designed for CPET and previously assessed in healthy adult populations (Green et al., 2023). Consists of 9 stages with gradual increments in speed and incline (see Table 1). The test consists of progressively increasing treadmill speeds and inclines over two-minute stages until voluntary exhaustion or when maximal heart rate has been achieved. The initial stage begins at a low workload, and each subsequent stage increases the speed (mph) and incline (%). For the experiment, in a stepwise ramp fashion, treadmill walking/ running will be performed. The protocol starts at 5 mph (1.5 % incline) and increases by 1 mph every two minutes, maintaining 1.5% incline during the first five stages. See Table 1 for further ramps in the experiment. HR and RPE will be measured at each stage to confirm equal physiologic and perceived workload during conditions. After the conclusion of the CPET, the participant will have a light 1-minute cool down and then sit on a chair for 5 minutes to recover baseline physiological variables. During the cool-down and sitting, the aforementioned values will be measured.

**Table 1.**
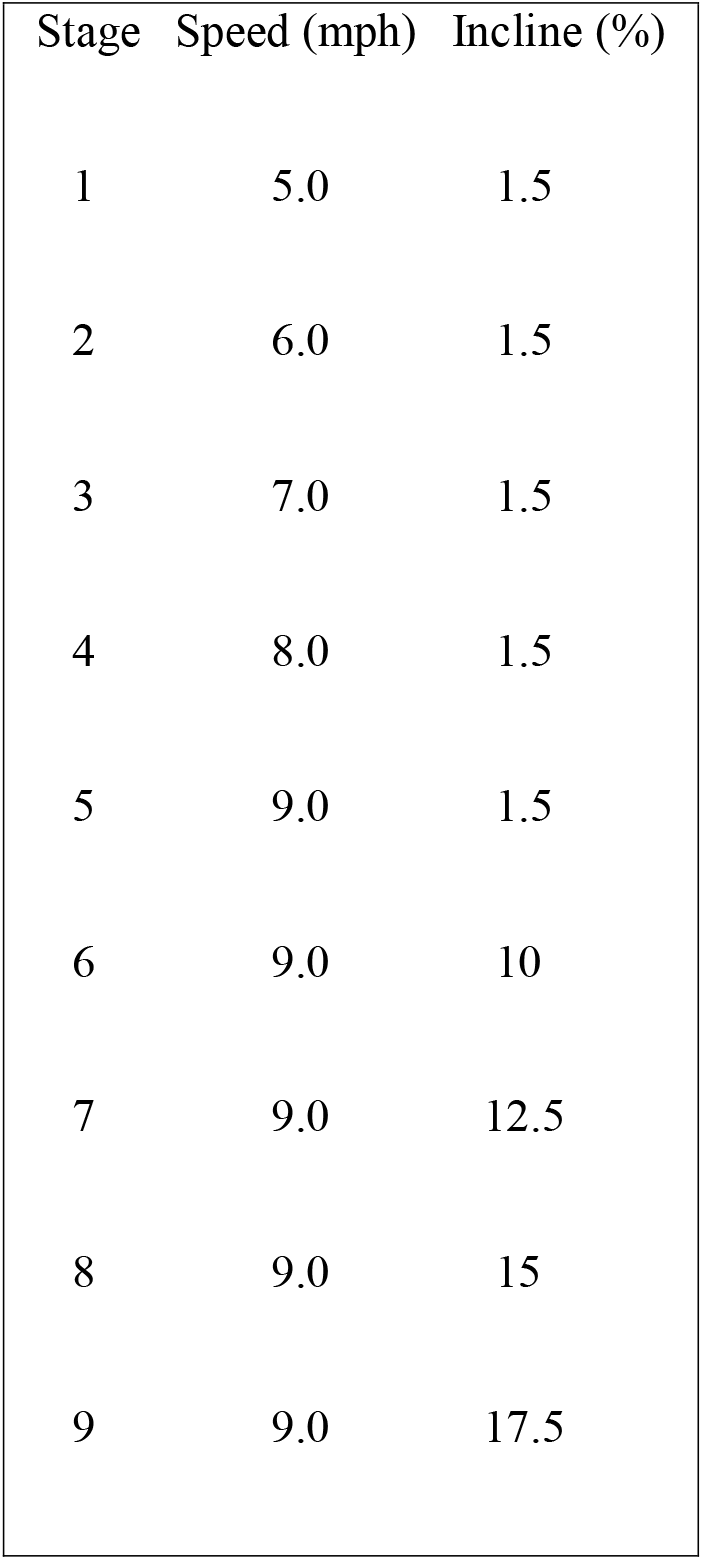
Treadmill test protocol stages, speed, and inclination for every two minutes.

Exercise intensity will be determined by using the Gellish et al. (2007) predicted HR maximum (HRmax) formula: HRmax = 207− (0.7 x age). This formula was selected because it is more accurate in the prediction of HRmax in well-trained individuals (Gellish et al., 2007; Cleary et al., 2011). A participant’s VO□max will be reached when one of the following conditions described by ACSM’s guidelines in exercise testing (ACSM, 2021) is met: a) an RPE between 9 to 10; b) HRmax reached as determined by the predicted Gellish et al. (2017) formula; c) final respiratory exchange ratio of 1.1 d) steady state of VO□ despite increasing intensity. We will measure the total time of exhaustion of the test and the final speed and incline corresponding to the last minute of the test.

#### Traditional walking warmup protocol

The WW will consist of 6 minutes of light to moderate treadmill walking at a comfortable pace with a 0% incline.

#### Suspension training dynamic warmup protocol

The second warmup will consist of using TRX equipment in a higher intensity circuit fashion of 6 minutes total. Table 2 shows the warm-up order and selection of exercises and duration time.

**Table 2.** Dynamic warm-up exercises with the suspension trainer TRX.

| Order | Suspension training exercise* | Duration (seconds) |
| --- | --- | --- |
| 1 | TRX Reverse Lunges | 45 |
| 2 | TRX Squats | 60 |
| 3 | TRX Jump Squats | 45 |
| 4 | TRC 45-degree rows | 30 |
| 5 | TRX 45-degree push-ups | 30 |

*\*Each exercise is followed by 30 seconds of rest.*

#### Outcome Measure

All anthropometric variables will be measured following standardized procedures as described by the American College of Sports Medicine (ACSM, 2021). Height will be measured using a wall-mounted stadiometer, and body mass will be measured using an electronic OMRON scale. Temperature will be measured with an electronic thermometer (Microlife; Taiwan). Blood pressure will be assessed by an automated sphygmomanometer. The RPE will be assessed using the Borg Scale (1-10) (Borg, 1982). Resting HR, resting BP, resting SPO□, resting temperature, and RPE will be taken at the beginning of the session in a seated position as baseline measurements of resting values. After the warm-up, immediately after the CPET protocol, and after 5 min of rest, these vital values will be retaken.

A calibrated metabolic system (VO□ Master Pro Systems, Vernon, Canada) will be used to assess gas exchange. The VO□ Master Pro is a portable device that operates through breathebyebreath gas exchange analysis. This analyzer is composed of a differential pressure flow sensor, an Oe analyzer (galvanic fuel cell sensor), which measures V□_E_ (large: 40 to 220□L·min−1, medium: 30 to 160□L·min−1, and resting: 5 to 40□L·min−1). The VO□ Master Pro system will be calibrated before use with a one□point calibration that uses room air and a 3□L syringe to calibrate the O□ and flow sensors. All metabolic variables will be collected via Bluetooth to an iPad tablet (Apple, Inc., Cupertino, CA, USA) equipped with the VO□Master Pro mobile app (Vernon, Canada, vo2master.com) for storage and later download of data. A Polar heart rate monitor/strap (Polar Electro Oy, Kempele, Finland) is integrated into the V0_2_ Master unit to record HR continually. The portable Bluetooth system allows users to perform suspension dynamic exercises without being impaired by cables. Participants will wear the VO□ Master Pro system during both trial conditions. The V0_2_ Master has previously shown acceptable validity and test-reliability for ventilatory assessment (Montoye et al., 2020, Thiessen et al., 2025).

### Statistical Methods

VO_2_ Master Pro data will be reintegrated to 30-second intervals for analysis and exported to Microsoft Excel. Descriptive statistics (mean□±□SD) will be reported. Primary outcomes will be analyzed using a linear mixed-effects model (LMM) with Order (day 1 vs day 2) as fixed effects and participant as a random intercept. Normality of paired differences will be assessed with the Shapiro-Wilk test; if normality is violated, a Wilcoxon signed□rank test will be used. Effect sizes will be reported as Cohen’s d (Cohen, 1988) for parametric tests or matched-pairs rank biserial correlation for non-parametric tests, with 95% confidence intervals. Equivalence testing of the two conditions will be done by two one□sided tests (TOST) analysis with a priori bounds of ±1 MET (±3.5 mL·kg□^1^·min□^1^) for VO□ and ±7 L·min□^1^ for V□_E_ as described in previous validation studies (Davis, 2024). Equivalence will be supported when both one-sided tests reach *p* < 0.05 and the 90% confidence intervals lie within the equivalence bounds. Analyses will be conducted in Jamovi (v2.3.21, Sydney, Australia) with the TOSTER module (v0.4.0) and the GaMLj (v2.8.0) module. Statistical significance will be set at α = 0.05.

## Data Availability

All data produced in the present study are available upon reasonable request to the authors

## Acknowledgements

Not applicable.

## Notes

### Competing Interest Statement

The authors have declared no competing interest.

### Clinical Trial

NCT07215052

### Funding Statement

Funded was provided by a Creativity and Research Grant by Monmouth University

### Author Declarations

Ethics/registration granted with number IRB SP2553 on June 25, 2025, by the Institutional Review Board of Monmouth University

